# National Validation of Risk Stratified Delivery Timing for NTSV Cesarean Reduction: A Population Based Analysis of 5.8 Million Deliveries

**DOI:** 10.64898/2026.03.31.26349855

**Authors:** Lauren Crabtree, Ciprian Paul Gheorghe

## Abstract

**Objective:** To externally validate a risk stratified delivery timing model for nulliparous, term, singleton, vertex (NTSV) cesarean reduction using national data.

**Design:** Population based cohort study of NTSV births in US National Vital Statistics System (NVSS) natality files, 2020–2024, using logistic regression for cesarean predictors and risk stratified Monte Carlo simulation (10,000 iterations per strategy and risk group) to evaluate delivery timing policies.

**Setting:** All live births in the US recorded in the NVSS natality files.

**Participants:** NTSV patients with term (37+ weeks) pregnancies and complete gestational age and delivery mode data (N=5 776 412). A sensitivity cohort excluded pre 39 week deliveries and pregnancies with preexisting diabetes or hypertension.

**Exposures:** Delivery timing strategies defined by gestational age and labor onset (elective induction at 39, 40, or 41 weeks, or expectant management to 42 weeks), evaluated within maternal age and body mass index (BMI) risk strata (low: age <35 and BMI <30; moderate: age ≥35 or BMI ≥30; high: age ≥35 and BMI ≥35).

**Main Outcomes and Measures:** Primary outcome was cesarean delivery, measured as the proportion of deliveries completed by cesarean across gestational ages, labor onset types, and age BMI strata. Secondary outcomes included gestational age specific cesarean rates, area under the receiver operating characteristic curve (AUC) for cesarean prediction, and simulated mean cesarean rates with 95% simulation intervals under four delivery timing strategies within each risk group.

**Results:** The overall NTSV cesarean rate was 26.4%. Cesarean Rates were U-shaped across gestational ages, with the lowest rate at 38 weeks (24.9%) and higher rates at 37 weeks (29.8%) and 41–42 weeks (28.1–28.5%). Risk group distribution was 64.9% low, 33.7% moderate, and 1.4% high. Model AUC was 0.65. Induction had higher cesarean rates than spontaneous labor (29.3% vs 24.2%; odds ratio 1.30, 95% confidence interval 1.29-1.30). Monte Carlo simulation favored induction at 39 weeks for high-risk patients (59.3%) and expectant management to 41–42 weeks for low-risk patients (19.1%).

**Conclusions and Relevance:** A risk stratified NTSV labor management model showed external validity in 5.8 million US births and consistently identified risk-specific timing strategies that lowered cesarean rates, supporting individualized delivery timing policies.

**Key Points:** *Question:* Among nulliparous, term, singleton, vertex (NTSV) pregnancies, can a previously developed risk stratified delivery timing model identify gestational age and induction strategies that reduce cesarean delivery when applied to US national data?

*Findings:* In this population based retrospective cohort study of 5.8 million NTSV births, external validation showed a U-shaped pattern in cesarean rates across gestational age and indicated that 39 weeks induction minimized cesarean risk for high-risk patients, whereas expectant management to 41–42 weeks minimized risk for low-risk patients.

*Meaning:* This risk stratified delivery timing model reliably identified strategies with lower cesarean delivery rates, supporting its use as a practical framework for counseling and institutional policy.

## INTRODUCTION

Cesarean delivery is one of the most common major operations performed worldwide and a central target of obstetric quality and safety initiatives.^1–2^ Efforts to reduce primary cesarean birth have led to benchmarks for low risk term pregnancies and widespread adoption of performance metrics that reward lower cesarean rates, even as more patients begin pregnancy at older ages, with higher body mass index (BMI), and with medical comorbidities that increase the risks of both cesarean delivery and prolonged labor.^3–6^ These changes raise concern that uniform cesarean targets and reduction strategies may not be appropriate or safe for all patients.^7–10^

Policy and practice efforts increasingly focus on the management of labor at term, particularly whether and when to offer induction of labor (IOL).^11–14^ Randomized trials show that elective induction at 39 weeks can lower cesarean rates among carefully selected low risk nulliparous patients without increasing short term perinatal morbidity, and observational data suggest modest population level declines in cesarean delivery as induction at 39 weeks has become more common.^11,15^ These findings have prompted some health systems to consider routine induction at 39 weeks as a strategy to improve cesarean metrics, despite uncertainty about whether similar benefits extend to more complex patient populations.

Emerging institutional analyses suggest that maternal age, pre-pregnancy BMI, gestational age at delivery, and induction status jointly shape cesarean risk, and that the delivery timing that minimizes cesarean risk may differ across maternal risk profiles.^16^ Simulation studies based on these data indicate that a single induction policy could lower cesarean risk for some groups while increasing it for others, but prior work has been limited to single centers or health systems without national representation or detailed neonatal outcomes.^16^

Whether similar risk dependent relationships among maternal characteristics, delivery timing, and cesarean risk are present at the national level remains unknown, limiting the development of delivery timing strategies and performance metrics that are both safe and robust across diverse practice settings. To address this gap, we used the National Vital Statistics System (NVSS) natality data from 2020 through 2024 to construct a contemporary national cohort of nulliparous term singleton vertex (NTSV) births.^17^ In this cohort, we applied and externally validated a previously developed risk stratification tool that integrates maternal characteristics and labor management strategies to estimate cesarean risk.^16^ We then evaluated how well this tool performs at the population level and whether delivery timing strategies informed by its risk strata may outperform universal timing approaches with respect to cesarean delivery.

## METHODS

### Study Design and Data Source

This population based cohort study included all NTSV deliveries recorded in the Centers for Disease Control and Prevention (CDC) NVSS natality detail files from 2020 through 2024. These files include individual-level birth certificate data for all registered live births in the United States based on the 2003 revision of the U.S. Standard Certificate of Live Birth. The data are publicly available and deidentified; therefore, institutional review board approval was not required.^17^

### Study Population

The NTSV cohort was defined according to Joint Commission PC-02 specifications^2^ as nulliparous patients delivering a live singleton in vertex presentation at 37 0/7 through 42 6/7 weeks of gestation. The primary cohort, henceforth referred to as the overall cohort, included all such deliveries with complete data on gestational age and mode of delivery (N = 5,776,412). A sensitivity cohort (n = 4,046,170) was limited to deliveries at 39 weeks or later and excluded pregnancies affected by preexisting diabetes or chronic hypertension, consistent with the sensitivity analysis performed in the index study.^16^

### Variables

The primary outcome was cesarean delivery, identified from the delivery method variable on the birth certificate. Gestational age was based on the obstetric estimate. Labor induction was classified from the IOL checkbox. Pre-pregnancy BMI was calculated from maternal height and pre-pregnancy weight as recorded on the birth certificate. Maternal age and source of payment were obtained as recorded. Race and ethnicity were based on patient self-report using predefined categories on the birth certificate, in accordance with current guidance for reporting these variables.

### Risk Stratification

Risk strata were defined as in the index study.^16^ The low-risk group included patients younger than 35 years with BMI less than 30. The moderate-risk group included patients who met one elevated risk criterion: age 35 years or older, or BMI 30.0 or higher. The high-risk group included patients who met both criteria: age 35 years or older and BMI 35.0 or higher. Patients with missing BMI data were excluded from risk stratified analyses but were retained in overall cohort analyses.

### Statistical Analysis

Cesarean rates were calculated by gestational age, labor onset type, and risk stratum with Wilson 95% confidence intervals (CI). The association between induction and cesarean delivery was estimated as an odds ratio (OR) with 95% CI. Logistic regression was used to evaluate maternal age, pre-pregnancy BMI, gestational age, and labor induction as predictors of cesarean delivery; model discrimination was assessed by the area under the receiver operating characteristic curve (AUC). A Monte Carlo simulation evaluated four delivery timing strategies within each risk group: elective induction at 39, 40, or 41 weeks, and expectant management to 42 weeks. For each strategy and risk group, 10,000 iterations were simulated using gestational age specific cesarean probabilities derived empirically from the corresponding risk stratum. The primary outcome was the simulated mean cesarean rate under each strategy, with 95% simulation intervals. Temporal trends in NTSV cesarean and induction rates were examined by year. Analyses were performed in Python (version 3.10). Anthropic Claude (Anthropic) assisted with generating Python code; authors verified all code and results.

## RESULTS

### Cohort Characteristics

Among 18,209,300 births across 2020–2024, 5,778,178 met NTSV inclusion criteria (31.7%). After exclusion of 1,766 records with incomplete data, 5,776,412 deliveries included the analytic cohort (Table 1). Mean maternal age was 27.3 years (SD 5.8), and mean pre-pregnancy BMI was 26.9 (SD 6.6). The induction rate was 43.2%. The overall NTSV cesarean rate was 26.4% (1,525,454/5,776,412). The sensitivity cohort comprised 4,046,170 deliveries with a cesarean rate of 25.9%.

**Table 1.** Baseline Characteristics and Outcomes of the Overall and Sensitivity Cohorts.

| Characteristic | Overall Cohort<br>(N=5,776,412) | Sensitivity Cohort<br>(n=4,046,170) |
| --- | --- | --- |
| Maternal age, y (SD) | 27.3 (5.8) | 27.3 (5.7) |
| Pre-pregnancy BMI (SD) | 26.9 (6.6) | 26.6 (6.3) |
| Induction of labor, % | 43.2 | 43.2 |
| Cesarean delivery, % | 26.4 | 25.9 |
| Risk group, % |  |  |
| Low risk | 64.9 | 66.9 |
| Moderate risk | 33.7 | 32.1 |
| High risk | 1.4 | 1 |

*Sensitivity cohort excludes patients with preexisting hypertension, diabetes and deliveries before 39 weeks.*

### Gestational Age and Cesarean Delivery

Cesarean delivery rates varied by gestational age in a U-shaped pattern (Figure 1). In the overall cohort, the lowest rate occurred at 38 weeks (24.9%), with higher rates at 37 weeks (29.8%) and at 41 and 42 weeks (28.5% and 28.1%, respectively). In the sensitivity cohort, cesarean rates were lowest at 39 weeks (25.5%) and increased at later gestational ages. At each gestational age, cesarean rates for induced labor exceeded those for spontaneous labor; at 40 weeks, induced labor was associated with a rate of 29.6% compared with 22.3% for spontaneous labor.

**Figure 1.**
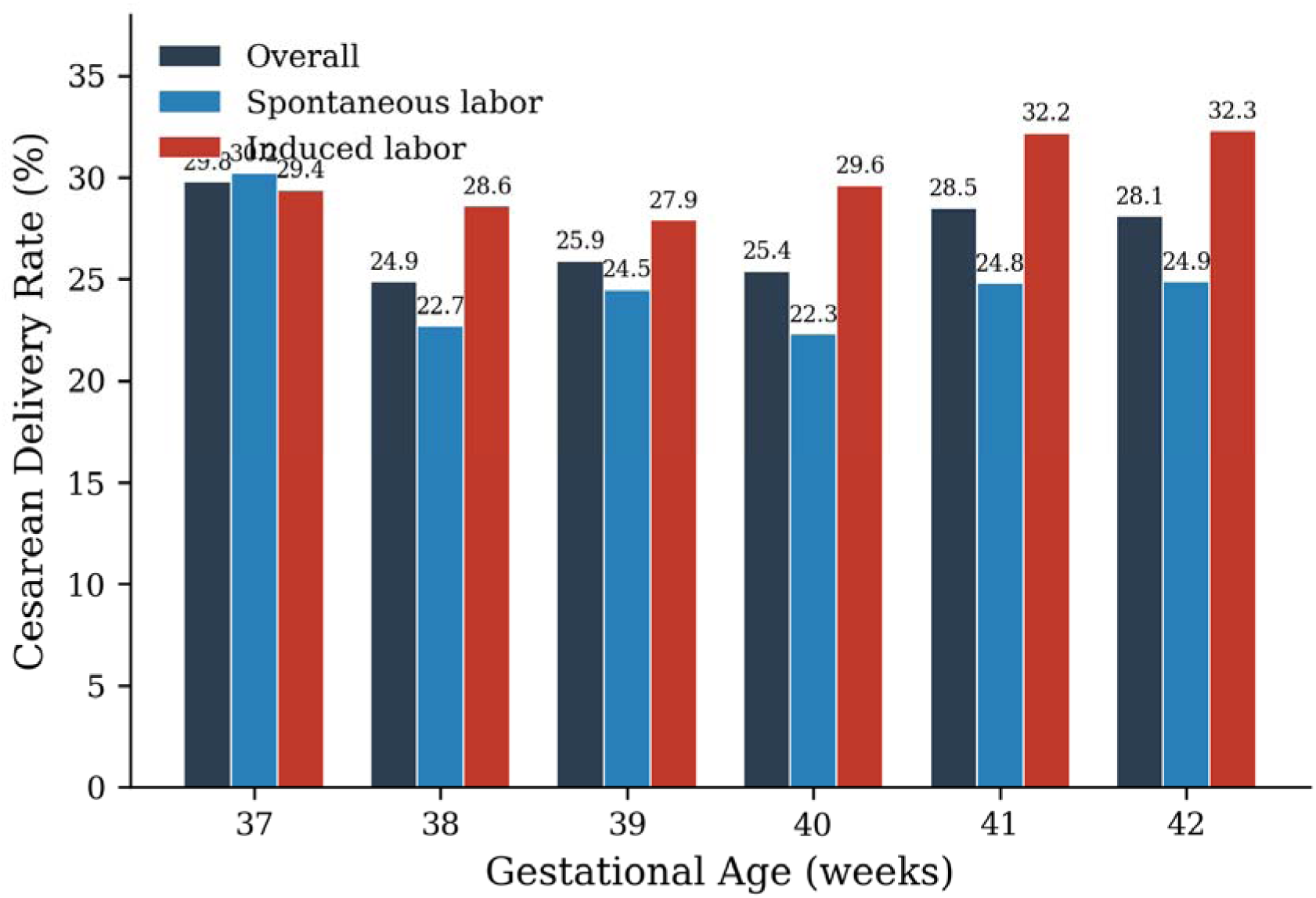
Cesarean delivery rates by gestational age and labor onset type in the overall NTSV cohort (N=5,776,412). Rates follow a U-shaped pattern with the nadir at 38 weeks. At each gestational age, induced labor was associated with higher cesarean rates than spontaneous labor.

### Predictive Model

Logistic regression identified maternal age (OR 1.527), pre-pregnancy BMI (OR 1.578), gestational age (OR 1.303), and labor induction (OR 1.182) as predictors of cesarean delivery. Model AUC was 0.647, indicating modest discrimination consistent with the multifactorial nature of cesarean decision-making and concordant with the AUC of 0.65 reported in the index study.^16^ The model was used to confirm the relative importance of these predictors and to support subsequent risk stratified simulation analyses, rather than to provide individualized bedside prediction.

### Labor Induction and Cesarean Delivery

In the overall cohort, cesarean rates were higher among induced compared with spontaneous labor (29.3% vs 24.2%; crude OR 1.30, 95% CI 1.29–1.30). Notably, the dataset did not distinguish elective from indicated induction, and these associations likely reflect residual confounding by indication.

### Risk Stratified Analysis

Among patients with complete BMI data, 64.9% were classified as low risk (n=3,684,620), 33.7% as moderate risk (n=1,914,091), and 1.4% as high risk (n=77,609) paralleling the distribution reported in the index study (62.4%, 33.9%, and 3.7%, respectively).^16^ Cesarean delivery rates increased across risk strata: 20.5% in the low-risk group, 36.5% in the moderate-risk group, and 57.9% in the high-risk group. As in the overall cohort, cesarean delivery rates were higher with IOL than with spontaneous labor in the low-risk (23.1% vs 18.8%; odds ratio [OR], 1.30) and moderate-risk (37.5% vs 35.6%; OR, 1.09) groups. In contrast, in the high-risk group, rates were lower with induction than with spontaneous labor (54.9% vs 61.9%; OR, 0.75).

The gestational age associated with the lowest cesarean rate was not uniform across strata (Figure 2). Among low-risk patients, the lowest cesarean rate for spontaneous labor occurred at 38 weeks (16.8%). In the moderate-risk group, the nadir for spontaneous labor was also at 38 weeks (34.4%). Among high-risk patients, cesarean rates for induced labor were lowest at 39 weeks (53.5%), replicating the index study finding that earlier planned delivery was associated with lower cesarean rates in this subgroup.

**Figure 2.**
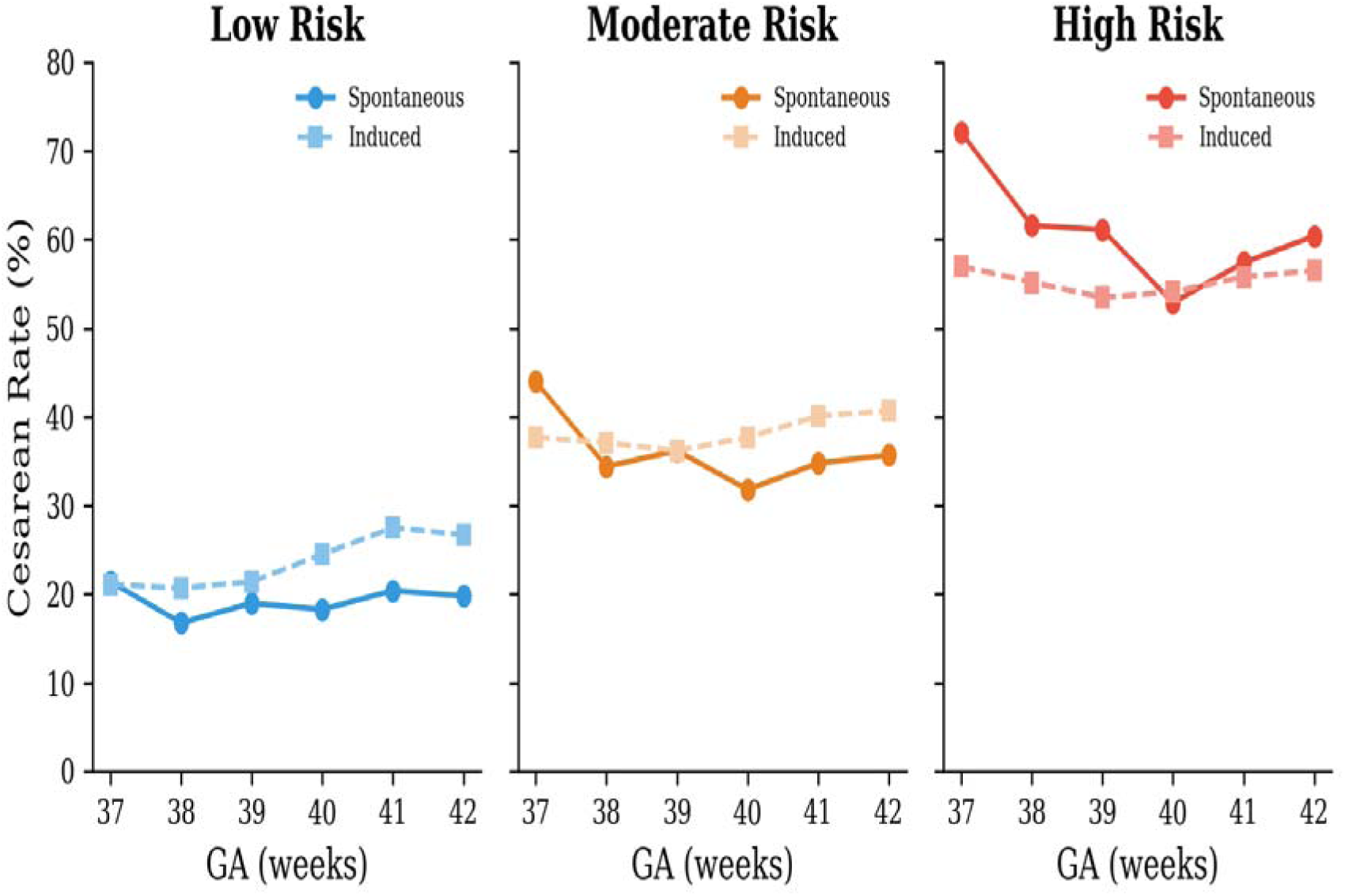
Risk-stratified cesarean delivery rates by gestational age and labor onset type. Among low-risk patients (left panel), cesarean rates for spontaneous labor are lowest at 38 weeks. Among high-risk patients (right panel), cesarean rates for induced labor are lowest at 39 weeks. Lines represent cesarean rates; circles, spontaneous labor; squares, induced labor.

### Monte Carlo Simulation

Simulation confirmed that no single induction timing minimized cesarean delivery across all risk strata (Table 2). In the high-risk group, induction at 39 weeks produced the lowest simulated cesarean rate (59.3%, 95% simulation interval [SI] 58.7–59.9%), with rates rising to 62% at 40 weeks. In the moderate-risk group, cesarean rates were lowest with 39-week induction (35.7%) compared with 40-week induction (37.1%). In the low-risk group, expectant management to 42 weeks yielded the lowest simulated cesarean rate (19.1%, 95% SI 19–19.2%), compared with 19.7% for 39-week induction.

**Table 2.** Monte Carlo Simulation: Simulated Cesarean Rates by Risk Group and Delivery.

| Strategy | Low Risk, %<br>(95% SI) | Moderate Risk,<br>% (95% SI) | High Risk, %<br>(95% SI) |
| --- | --- | --- | --- |
| Induction at 39 weeks | 19.7 (19.6–19.7) | 35.7 (35.6–35.8) | 59.3 (58.7–59.9) |
| Induction at 40 weeks | 20.5 (20.4–20.6) | 37.1 (37–37.2) | 62 (61.4–62.6) |
| Induction at 41 weeks | 19.7 (19.6–19.7) | 36.2 (36.1–36.4) | 61.7 (61.1–62.3) |
| Expectant to 42 weeks | 19.1 (19–19.2) | 35.9 (35.7–36) | 61.7 (61.1–62.3) |

*SI, simulation interval. Bold values indicate the lowest simulated cesarean rate within each risk group.*

### Temporal Trends

Over the 5 year study period, NTSV cesarean rates increased from 25.9% in 2020 to 26.7% in 2024. In parallel, induction rates rose from 41.5% to 45.1%.

## DISCUSSION

In this analysis of 5,776,412 NTSV deliveries, a risk stratified delivery timing model for NTSV deliveries originally developed in a single tertiary center showed robust external validity at the national level. The U shaped association between gestational age at delivery and cesarean birth, the predictive importance of maternal age and pre-pregnancy BMI, and the risk dependent differences in optimal induction timing initially demonstrated in a single center cohort of 10,525 deliveries were all confirmed at the national level with greater precision.^16^ These findings support the use of a structured, age and BMI based framework to guide NTSV induction decisions in clinical practice, as illustrated in Figure 3.

**Figure 3.**
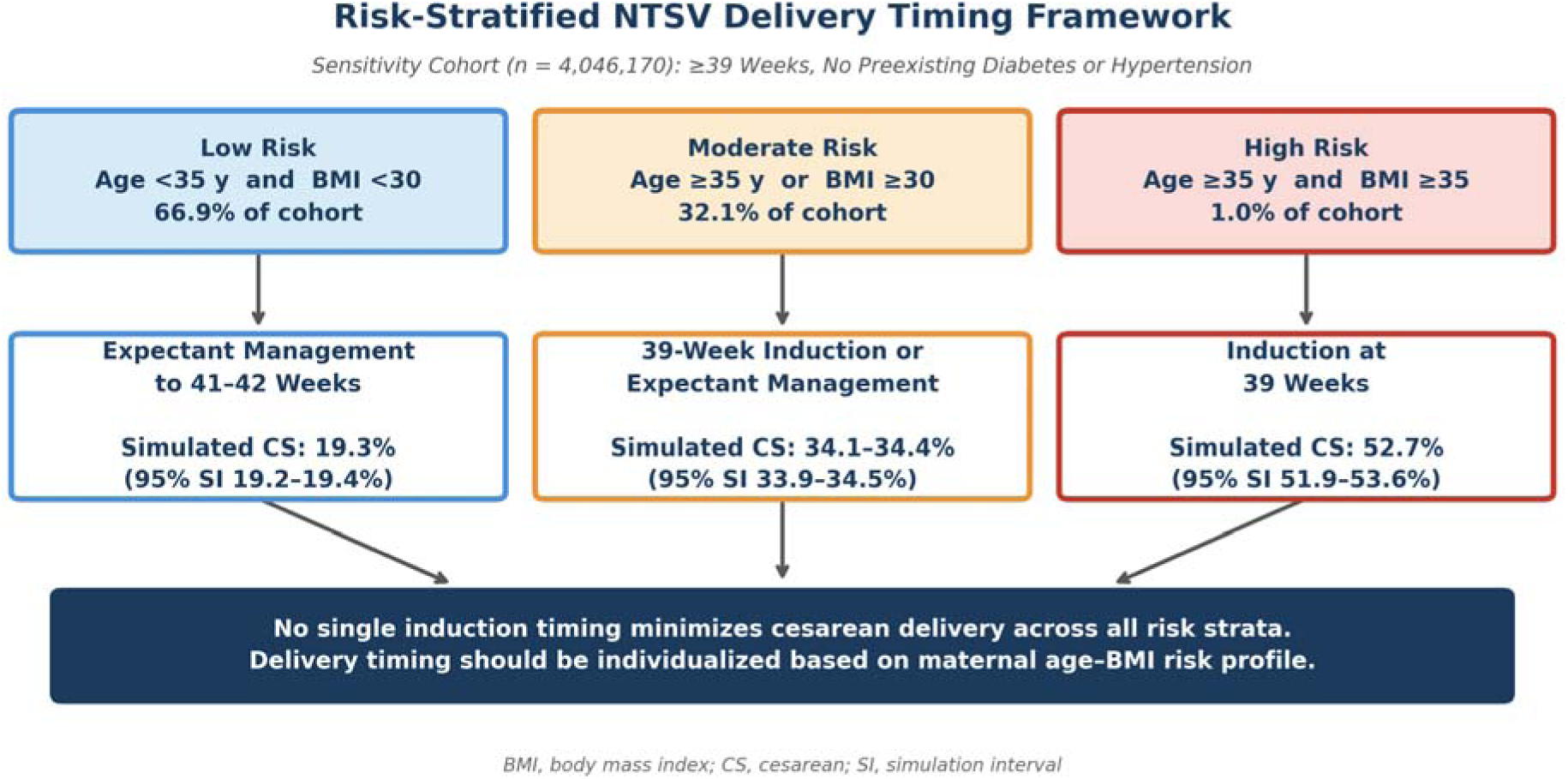
Decision framework for risk stratified delivery timing in NTSV pregnancies, derived from Monte Carlo simulation in the sensitivity cohort (n = 4,046,170). Patients are classified into three risk strata based on maternal age and pre-pregnancy body mass index. Optimal delivery timing strategy and corresponding simulated cesarean rates with 95% simulation intervals are shown for each stratum. No single induction timing minimized cesarean delivery across all strata; optimal timing differed by maternal risk profile. BMI, body mass index; CS, cesarean delivery; SI, simulation interval.

A key observation is the U shaped relationship between gestational age and cesarean delivery in the overall national cohort, with higher cesarean rates at both earlier and later term gestations and lower rates in the mid term range. Cesarean risk also varied by maternal age and pre-pregnancy BMI, with higher absolute cesarean rates in the moderate and high risk groups at nearly every gestational age compared with the low risk group. Together, these patterns indicate that baseline maternal risk and gestational age each make independent contributions to cesarean risk, supporting delivery timing strategies that incorporate both dimensions rather than a single gestational age threshold for all NTSV patients.

The Monte Carlo simulations further demonstrated that the association between elective induction and cesarean delivery is strongly dependent on baseline maternal risk and gestational age. Among patients at highest baseline risk, earlier planned delivery around 39 weeks gestation produced the lowest simulated cesarean rates, consistent with the interpretation that these patients accumulate cesarean risk with advancing gestation more rapidly than the risk introduced by induction itself. In contrast, among low- and moderate-risk patients, later induction or continued expectant management to 41–42 weeks was associated with lower simulated cesarean rates than routine induction at 39 weeks. Collectively, these simulations confirm the index study’s finding that no single induction timing minimizes cesarean delivery across all risk strata and that the gestational age at which cesarean risk is lowest differs meaningfully by maternal risk profile.^16^

These findings should be interpreted alongside randomized trial data on induction at 39 weeks.^11^ The ARRIVE Trial demonstrated reduced cesarean delivery with induction at 39 weeks among carefully selected low risk nulliparous patients under randomized conditions, whereas our analysis evaluated all contemporary NTSV births in the United States across the full spectrum of maternal age, BMI, and comorbidity.^11,16^ While our observational design cannot replicate the intent-to-treat framework of ARRIVE, and the inability to distinguish elective from indicated induction represents a fundamental difference in study populations that limits direct comparison, the temporal trends observed in the present analysis (a concurrent rise in both induction rates (41.5% to 45.1%) and NTSV cesarean rates (25.9% to 26.7%) from 2020 to 2024) suggest that increasing induction utilization at the population level has not yielded the reduction in cesarean deliveries anticipated from trial data. In this light, these observations are consistent with the hypothesis that real-world induction practices, which include a heterogeneous mix of indications and patient risk profiles, may produce net effects that differ from those observed under controlled conditions.

This study has several strengths. Use of the complete US natality data over a 5 year period yielded a national cohort of 5.8 million NTSV deliveries, eliminating small cell sizes at gestational age extremes and allowing precise estimation of cesarean probabilities across gestational ages and maternal risk strata. Applying the same age and BMI based risk stratification framework as the index study enabled direct comparison between institutional and population level findings and demonstrated that the model’s structure generalizes across diverse practice settings.^16^ The combination of empirical risk estimates and Monte Carlo simulation permitted evaluation of plausible delivery timing strategies at a policy level that would be difficult to test directly in randomized trials.

However, important limitations should also be acknowledged. Birth certificate data lack granularity on cervical status (Bishop score), indication for induction, labor progress, and indication for cesarean delivery, which precludes adjusted analyses that fully account for confounding by indication. In addition, prepregnancy BMI relies on self reported weight and may therefore be misclassified, although such error would likely attenuate rather than exaggerate observed risk gradients across BMI defined strata.

This analysis provides national external validation of a risk stratified framework for NTSV delivery timing, demonstrating that a model derived from a single center data generalizes to 5.8 million contemporary deliveries. Our findings indicate that optimal delivery timing varies meaningfully by maternal risk profile and that a universal 39 week induction policy does not minimize cesarean rates for all NTSV patients. Future work should extend this validated risk stratified delivery timing model to incorporate neonatal and other maternal outcomes, enabling explicit evaluation of maternal and neonatal trade offs across gestational ages and induction strategies and supporting implementation of risk stratified induction policies in diverse practice settings.

## Data Availability

All data produced in the present study are available upon reasonable request to the authors

## ACKNOWLEDGEMENTS

We acknowledge the National Center for Health Statistics and the state and local vital registration offices for collecting and providing access to the National Vital Statistics System natality data used in this study; the findings and conclusions in this report are those of the authors and do not necessarily represent the views of the National Center for Health Statistics.

